# Neural Changes to Transcranial Alternating Current Stimulation in the Gamma Range Over the Left Frontoparietal Network: A Preliminary eLORETA EEG Study

**DOI:** 10.1101/2025.01.24.25321077

**Authors:** Tien-Wen Lee, Gerald Tramontano

**Author notes:** Corresponding Author: Tien-Wen Lee.

## Abstract

Applying transcranial alternating current stimulation (tACS) at 40 Hz to the frontal and parietal regions can improve cognitive dysfunctions. This study aimed to explore the neural changes following transcranial electrical stimulation treatment. Electroencephalography (EEG) recordings were obtained from a cohort of 34 participants with various cognitive impairments before and after 12 sessions of tACS treatment. Alternating currents at 2.0 mA were administered to the electrode positions F3 and P3 for 20 minutes of each session, following the 10-20 EEG convention. Using eLORETA, scalp-recorded signals were reconstructed into cortical current source density (CSD). We then assessed the differences in power and connectivity strength across multiple spectra (delta to gamma). We observed a consistent trend of decreased CSD at the stimulating sites across different spectra, most prominent at beta and gamma bands (*P* < 0.01). On the contrary, the right hemisphere showed a trend of increased CSD, which was likely mediated by inter-hemispheric rivalry. In addition, the connectivity strength between the left frontal and parietal regions increased significantly (*P* = 0.017). Artificial injections of tACS would de-synchronize regional oscillation and enhance inter-regional crosstalk. The pattern of neural changes was concordant with our previous tACS reports (5-Hz), suggesting common neural mechanisms driving the neurophysiological effects of tACS.

## Introduction

Cortical oscillations at various frequencies have been linked to diverse cognitive functions (Klimesch et al., 1996; Klimesch, 1999; Lachaux et al., 2012). For instance, working memory and semantic memory functions have been respectively associated with power changes in the theta and upper alpha bands (Klimesch, 1999; Brzezicka et al., 2019). Gamma rhythms have been linked to the functioning of extensive brain networks and cognitive processes such as attention, perceptual grouping, and working memory (Brookes et al., 2011; Fell et al., 2002; Lundqvist et al., 2018). With the introduction of transcranial alternating current stimulation (tACS), it becomes possible to inject artificial oscillation into the brain and study its influence on neural rhythmicity and cognition (Antal et al., 2008; Bland and Sale, 2019; Ali et al., 2013).

Among the physiologically pertinent frequency bands of brain waves, we are particularly interested in gamma, given its coupling with downward spectra (delta to beta), co-bursts with beta in volitional control, and its roles in cognitive impairment in various neurocognitive conditions (Foster and Parvizi, 2012; Goodman et al., 2018; Belluscio et al., 2012; Canolty et al., 2006; Gągol et al., 2018; Lundqvist et al., 2018; Grigorovsky et al., 2020; Gong et al., 2021; Bonnefond and Jensen, 2015; Rossini et al., 2006; Moretti et al., 2012). Although transcranial electrical stimulation (tES) has been widely applied as a tool for basic and clinical research (Guleyupoglu et al., 2013), studies have supported tACS as a treatment option for neuropsychiatric conditions (Clancy et al., 2018; Alexander et al., 2019; T. W. Lee et al., 2024), such as depression and anxiety. Gamma tACS has been shown to improve the performance of working memory and fluid intelligence (Hoy et al., 2015; Santarnecchi et al., 2016; Booth et al., 2022). A sporadic report suggested that tACS at 40 Hz was promising in the prevention and treatment of Alzheimer’s disease (McDermott et al., 2018). Our recent clinical study delivered 40 Hz tACS to cognitively impaired populations and confirmed its efficacy and potential in clinical practice (T. W. Lee and Tramontano, 2024a).

Despite its increasing popularity, the neural mechanisms underlying the therapeutic efficacy of tACS remain unclear. It is verified that tACS may change the firing rate and spike timing of neuronal activities in a spectra-specific manner (Anastassiou et al., 2010; Radman et al., 2007; Radman et al., 2009; Krause et al., 2019). It was thus inferred that applying a particular frequency of tACS can “synchronize” the neural oscillations to match the frequency of the electrical stimulation, framed as “entrainment theory” (Zaehle et al., 2010; Antal and Paulus, 2013). The evidence of tACS-induced entrainment in the human brain dynamics, however, has been controversial (Helfrich et al., 2014; Voss et al., 2014; Brignani et al., 2013; Alexander et al., 2019; Lafon et al., 2017). Our previous EEG research of 5-Hz tACS over the right hemisphere has provided novel insight into this pivotal issue (T. W. Lee and Tramontano, 2024b; c). By comparing the EEG collected before and after 25 min tACS at 5-Hz (in theta range), we observed spectrum-specific and -unspecific alterations, summarized below. First, tACS at 5 Hz desynchronized regional theta power and enhanced contralateral theta power. The latter was likely mediated by the neural mechanism of inter-hemispheric rivalry (Kinsbourne, 1974; Hilgetag et al., 2001; Naeser et al., 2005). The areas undergoing modulation were widespread, consistent with tES being a broad-spectrum technique, as opposed to the more focal transcranial magnetic stimulation. Second, tACS imposed wide-spread cross-spectral influence on power, well beyond the “entrained” frequency—spectral-unspecific. Last, inter-regional connectivity (phase synchronization) increased in a spectrum-specific way, which only occurred at the theta range.

Continuing from our previous work, this research explored the neural consequences of 40-Hz gamma tACS applied over the left frontoparietal network across 12 treatment sessions, aiming to enhance cognitive capability. Both regional power and inter-regional connectivity profiles were examined. In line with the brain-based approach, we utilized exact low-resolution brain electromagnetic tomography (eLORETA) to map EEG data from the scalp onto the gray matter voxels of a standardized brain template, i.e., current source density (CSD) (Roberto D Pascual-Marqui, 2007a; Jurcak et al., 2007). The power changes of CSD following treatment were examined statistically. The connections between the frontal and parietal areas, characterized by different frequency spectra, were assessed using lagged coherence, which is a linear measure, and phase synchronization, a nonlinear indicator of functional connectivity (Roberto D Pascual-Marqui, 2007b). We predicted that: (1) the left and right frontoparietal networks showed opposite trends of power changes, with tACS exerting unfavorable influence on the underlying neural synchronization; (2) the power modulation was not limited to the gamma range, but extended to the lower frequency ranges, i.e., cross-spectral effects; (3) the functional connectivity strength increased between frontal and parietal regions. The influence of gamma tACS on working memory and executive control was evaluated using selected tasks from the Delis-Kaplan Executive Function System (D-KEFS) (Delis et al., 2001).

## Materials and Methods

### Participants and working memory assessment

This study focused on patients exhibiting cognitive deficits who underwent 40 Hz tACS treatment applied to the left frontal and parietal regions. We reviewed data collected from our clinics between 2018 and 2022, following prior approval of research protocol by the private review board Pearl IRB, with approval ID 2023-0133. To be included in this study, patients need to have a diagnosis characterized by cognitive deficits (e.g., mild cognitive impairment [MCI], learning disability, attention-deficit hyperactivity disorder [ADHD]) or have a chief complaint of cognitive decline. Trails A and B, letter fluency, and design fluency were selected to assess processing speed, cognitive flexibility, verbal fluency, and non-verbal fluency, respectively. Individuals with epilepsy, skull defects, intracranial electrodes, brain lesions, vascular clips or shunts in the brain, cardiac pacemakers, or other implanted biomedical devices, as well as those who are pregnant or lactating, were deemed ineligible for tES in accordance with safety guidelines (Matsumoto and Ugawa, 2017; Antal et al., 2017). Written informed consent was obtained from all clinical patients. For participants under 18 years of age, the consent was provided by their parents, legal guardians, or next of kin, in accordance with ethical guidelines requiring appropriate consent for minors. Participants were instructed to maintain their current treatments, including medications, without any dosage adjustments during the tES treatment. The primary focus of this research was the changes in EEG following tES. We identified 34 cognitively impaired patients who received full tACS treatment and underwent pre- and post-treatment EEG.

### Administration of tACS

We employed an Investigation Exempt Device (IDE), the Starstim-8 neuromodulation device, developed by Neuroelectrics, Inc. (Barcelona, Spain), to attenuate cognitive deficits arising from different causes. The device utilizes electrodes connected through wires to a rechargeable battery, delivering electric currents directly to the scalp and brain. Prior to attaching the electrodes, the scalp underwent a gentle cleansing with skin preparation gel. Subsequently, conductive gel was applied to facilitate proper electrode-to-scalp contact. These steps were essential to maintain optimal electrode contact and ensure that the impedance level remained below 5 k ohms (DaSilva et al., 2011).

The montage of electrodes covered the left lateral side of the head at F3 and P3 positions in terms of the 10-20 EEG convention. The peak current intensity was 2.0 mA, with sinewave currents oscillating at 40 Hz and alternating between electrodes F3 and P3, see Figure 1. During the neuromodulation session, the subjects were asked to practice computerized cognitive training games (https://www.happyneuronpro.com/). To familiarize patients with tES, we implemented a gradual dose escalation strategy during the initial three sessions. This strategy involved the following current levels: 1.0 mA for the first session, 1.5 mA for the second session, and 2.0 mA for the third session. Each stimulation session had a duration of 25 minutes, commencing with a 1-minute ramp-up phase and concluding with a 30-second ramp-down phase to minimize skin irritation. The 12 treatment sessions were completed in 3 weeks.

**Fig. 1.**
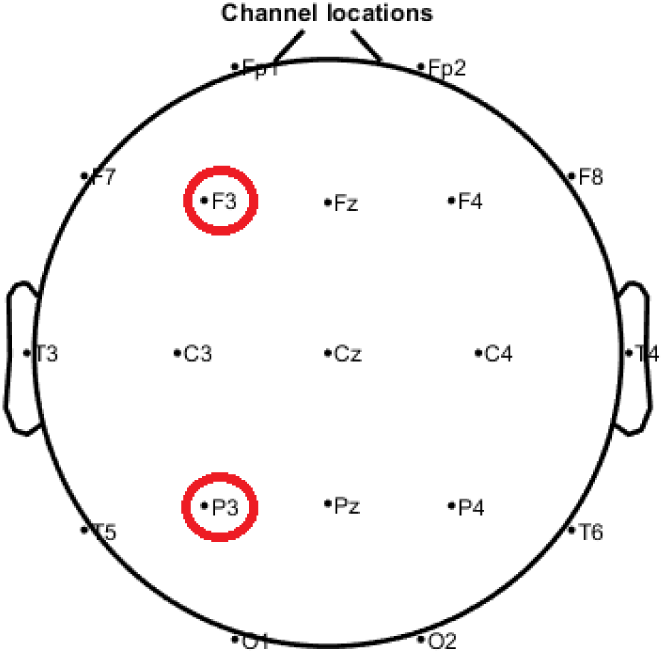
An illustration of the F3 and P3 positions on the scalp.

### EEG recording and data pre-processing

Before the initial treatment and after the last neuromodulation session, we employed the Brainmaster device (Discovery 24, with bandwidth 0 to 80 Hz, 19 channels; https://brainmaster.com/) to collect 10-minute EEG data with eyes open at a sampling rate of 256 samples per second, utilizing a linked-ear reference. EEG recording involved the following steps: Participants were seated comfortably, their scalp was cleaned, and a customized electrode cap was aligned according to the 10-20 system. Conductive gel was applied, and impedance was checked to ensure it remained below 5 ohms. Signal quality was verified and continuously monitored throughout the recording to ensure clarity and participant compliance. We utilized the EEGLAB software to edit the EEG traces (Delorme and Makeig, 2004). Data preprocessing included band-pass filtering (1–50 Hz), automatic artifact removal (Automatic Artifact Removal [AAR] toolbox for electromyographical activity mitigation and Artifact Subspace Reconstruction [ASR] toolbox for burst correction) (Gómez-Herrero et al., 2006; Blum et al., 2019), and manual elimination of the remaining noisy portions, including those linked to blinks, flutter, eye movements, line movement, sweat artifact and so on. The above-mentioned processing was conducted by a single rater (TW Lee). After eliminating artifacts, the cleaned EEG data were divided into 2-second epochs and then imported into eLORETA for further analysis.

### eLORETA and power analyses

The eLORETA method was employed to transform scalp EEG signals into neural electrical activities of 6,239 gray matter voxels of an MNI template (Roberto D Pascual-Marqui, 2007a; Jurcak et al., 2007). Unlike the conventional parametric approach of source localization, eLORETA utilizes a linear imaging method with adaptive data-dependent weights to account for measurement and biological noise. It accurately computes arbitrary point-test sources with precise, error-free localization. eLORETA, based on linearity and superposition principles, is well-suited for mapping distributed electric sources (CSD) in the brain cortex, albeit with limited spatial resolution. The power spectrum of delta (1–4 Hz), theta (4–8 Hz), alpha (8–12 Hz), beta (12–30 Hz), and gamma (30–50 Hz) for each voxel were derived (T. W. Lee and Tramontano, 2023).

We used paired t-tests and a non-parametric statistical method (SnPM) with 5,000 randomizations to compare cortical CSD powers before and after treatment (Westfall and Young, 1993; Nichols and Holmes, 2002). The max-statistics and corrected *P*-values for multiple testing can thus be derived. We employed the exceedance proportion test in eLORETA software to assess the significance of activity based on its spatial extent, identifying clusters of supra-threshold voxels. The null hypothesis was that there were no differences in CSD powers after tES treatment, with a two-tailed significance level of P < 0.05. A more detailed explanation of the principles behind eLORETA and non-parametric exceedance proportion test can be referred to our recent report and the source papers (Roberto D Pascual-Marqui, 2007a; Nichols and Holmes, 2002; T. W. Lee and Tramontano, 2024c).

### Connectivity analysis

We used lagged (general) coherence and phase synchronization to represent linear and nonlinear functional connectivity strength, respectively (Roberto D Pascual-Marqui, 2007b). Both were computed from the CSD time series of the selected voxel. The equations for phase synchronization are similar to those for coherence, except for a pre-normalization step that accounts for power influence, making it a measure of nonlinear connectivity unaffected by amplitude relationships. Total coherence comprises both lagged and instantaneous dependence. However, because the instantaneous part is influenced by non-physiological factors like volume conduction and low spatial resolution, it was omitted in this report (Roberto D Pascual-Marqui, 2007b). For the same reason, we only focused on the lagged component of phase synchronization.

The average connectivity strengths were calculated between two coordinates, which were the center of Brodmann area (BA) 8/9 (frontal; [-30,30,40]) and BA 39/40 (parietal; [-45,-50,40]). Paired t-tests were used to examine the connectivity changes before and after the tACS treatment course.

## Results

The mean age of the 34 patients ranged from 10.9 to 84.0 years, with a mean ± SD of 60.0 ± 24.7 years. Among them, there were 15 males and 19 females. All selected patients tolerated the maximum tES current at 2.0 mA. Their diagnoses include MCI (n=12), ADHD (10), mild to moderate intellectual disability (4), learning disability (6), and traumatic brain injury (2). Only mild tACS-related side effects, such as tingling or itchy, were observed. Changes in test scores were observed for Trail A, Trail B, and letter fluency, but not for design fluency; see Table 1.

**Table 1.** The comparative results of selected neuropsychological tests pre- and post-treatment (post - pre differences)

|  | Pre-Tx | Post-Tx | t-scores | p values |
| --- | --- | --- | --- | --- |
| **Trail A** | 40.8 (19.0) | 33.5 (14.0) | -2.85 | 0.0079 |
| **Trail B** | 116.9 (60.3) | 97.3 (42.9) | -3.48 | 0.0016 |
| **Letter Fluency** | 28.0 (10.7) | 31.3 (10.9) | 3.20 | 0.0033 |
| **Design Fluency** | 24.2 (8.6) | 26.9 (9.2) | 2.04 | 0.0510 |

Tx: treatment; subject number = 30.

### Power analysis and statistical comparisons

The neural responses to 40Hz tACS exhibited a consistent pattern across different spectra: (1) at the left frontoparietal region, the same side of tES, there was a significant reduction in CSD powers at gamma and beta frequencies. Both the peak t-values and the spatial extents surpassed the statistical thresholds, with the *P* values at the level of 0.005. Notably, the trend propagated to lower frequency bands; (2) at the right frontoparietal region, contralateral side of tES, there was a significant enhancement of CSD powers. The peak t-values did not survive the statistical challenge, but the spatial extent remained significant. Again, the observed trend extended to lower frequency bands. The results of power analyses are summarized in Table 2 and illustrated in Figures 2 and 3.

**Fig. 2.**
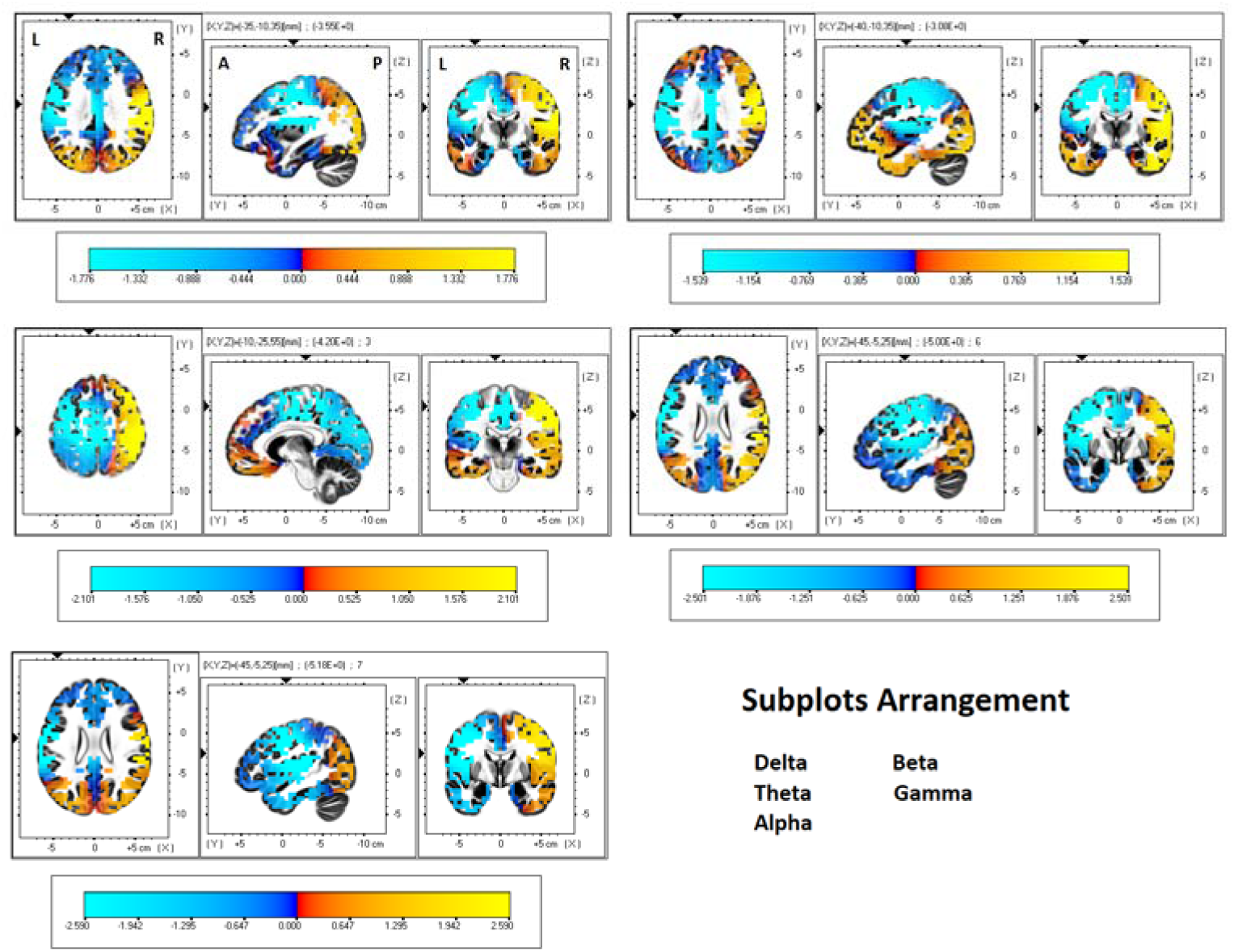
The changes in CSD powers (post-treatment minus pre-treatment) are represented at five defined spectra (delta, theta, alpha, beta, and gamma). Warm colors indicate an increase, while cool colors indicate a decrease. The axial (left), sagittal (middle), and coronal (right) sections are centered around the highest statistics. A color bar is included at the end of each sub-graph, scaled to 50% of the maximum to provide a clearer illustration of the trend in CSD changes for each frequency band.

**Fig. 3.**
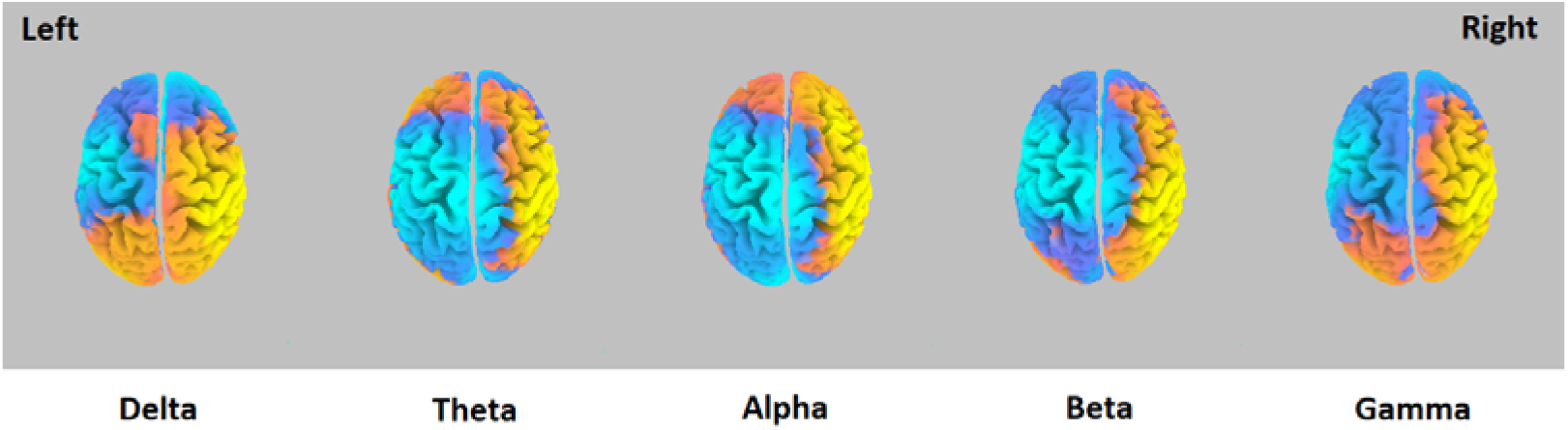
Top views of the CSD differences following tACS treatment, highlighting a consistent trend of power changes across different spectra. The color bars can be referred to in Figure 2.

**Table 2.**
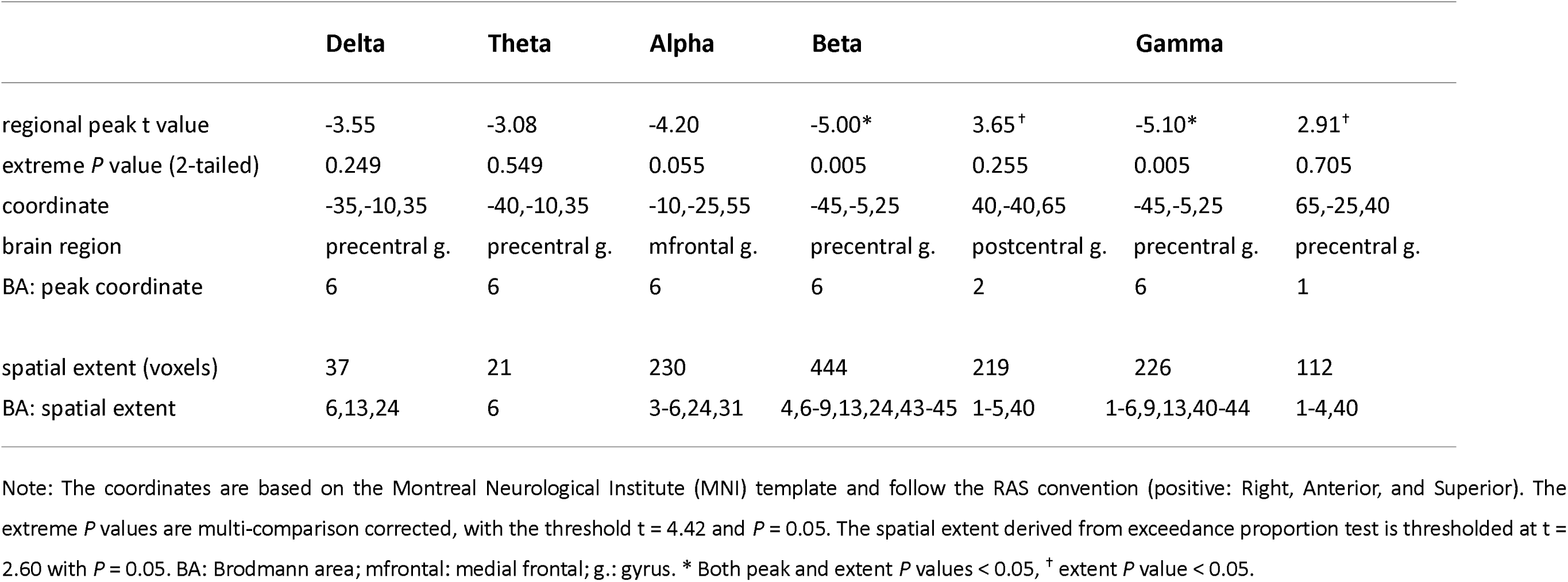
Summary of neural changes to neuromodulation treatment, with the points with the highest t values reported.

### Connectivity analysis and statistical comparisons

Our hypothesis was verified that the connectivity strengths were increased after the 40-Hz tACS over the frontoparietal network. The enhancement was statistically significant for phase synchronization, not coherence. The connectivity strengths and the statistics are summarized in Table 3, and the significant results are illustrated in Figure 4. Supplementary analyses showed that the connectivity changes were only present in the gamma range, not in other spectra (spectrum-specific; data thus not shown).

**Fig. 4.**
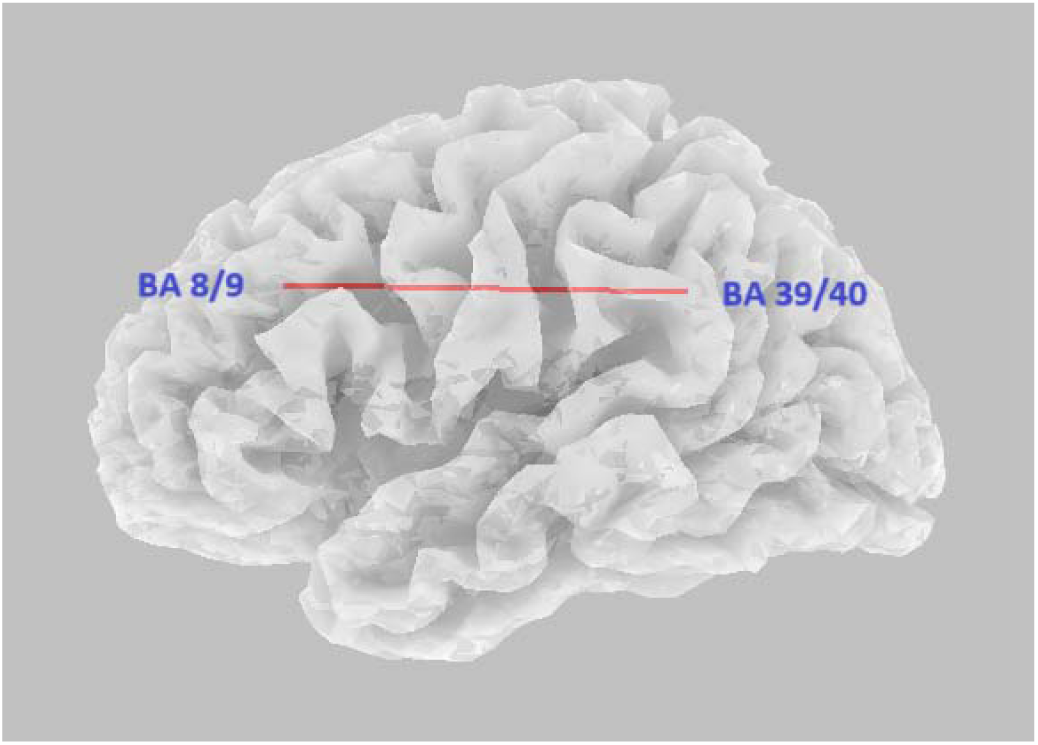
The lagged phase synchronization was increased in the frontoparietal network. Centers of Brodmann areas serving as neural nodes for connectivity analyses are labeled.

**Table 3.** Lagged general coherence and phase synchronization were computed between the central coordinates of BA 8/9 and BA 39/40 and were compared between post-treatment and baseline conditions.

|  | Mean (SD) |  | t score | P value |
| --- | --- | --- | --- | --- |
| **Lagged General Coherence** |  |  |  |  |
|  | Post-tACS | Baseline |  |  |
| **BA8/9 and BA39/40** | 0.0027 (0.002) | 0.0022 (0.002) | 1.05 | 0.302 |
| **Lagged Phase Synchronization** |  |  |  |  |
|  | Post-tACS | Baseline |  |  |
| **BA8/9 and BA39/40** | 0.0024 (0.002) | 0.0014 (0.001) | 2.50 | 0.018* |

\* P value < 0.05. BA: Brodmann Area.

## Discussion

Neural oscillation is a fundamental characteristic of living organisms (T. W. Lee, 2016), which endorses various cognitive functions and underlies abnormalities in neurocognitive disorders (McDermott et al., 2018; T. W. Lee and Tramontano, 2023). In addition to offering a research gateway to modulate brain rhythms and cognitive processes, tACS at the gamma range can improve clinical cognitive dysfunctions (Ali et al., 2013; Antal and Paulus, 2013; T. W. Lee and Tramontano, 2024a; Hoy et al., 2015; Santarnecchi et al., 2016; Booth et al., 2022). We utilized EEG to investigate neural alterations subsequent to 12 treatment sessions of 2mA tACS at 40 Hz over left frontal and parietal regions, which aimed to ameliorate cognitive deficits resulting from diverse diagnoses. Through the comparisons of post-treatment minus baseline neural indices, our three hypotheses were confirmed, aligning closely with the findings from our previous 5-Hz tACS and EEG studies that employed a similar experimental design and analytic strategy (T. W. Lee and Tramontano, 2024b; c). As both tES and eLORETA are low-resolution tools, we discussed the results on a lobar or hemispheric scale, instead of focusing on the peak coordinates, which were the typical approach in previous neuroimaging studies.

### Power spectra changes associated with tACS treatment

We noticed reduced CSD powers on the same side of alternating current application and increased powers on the opposite side of the brain. This pattern was most pronounced in the gamma and beta frequency ranges, and a similar trend was also observed in the lower frequency bands, from delta to alpha. The findings offer two important insights. First, artificial injection of alternating currents into the brain interfered with, rather than synchronized, underlying neural oscillation across spectra. Although increased neural oscillation to tACS had been inferred from animal studies and entrainment theory (mainly from real-time recording) (Anastassiou et al., 2010; Radman et al., 2007; Radman et al., 2009; Krause et al., 2019; Zaehle et al., 2010; Antal and Paulus, 2013), recent research has not consistently validated the prediction but has instead suggested alternative neural mechanisms in the offline conditions, such as plasticity. For example, application of alpha tACS targeting the dorsolateral prefrontal cortex decreased the alpha power (Alexander et al., 2019). Delivering tACS at 6 Hz and 10 Hz over the occipito-parietal area impaired performance in the detection task, indicating that an interference rather than facilitation of the underneath neural processing (Brignani et al., 2013). Lafton et al. conducted intracranial recordings and found no evidence of sleep rhythm entrainment induced by tACS (Lafon et al., 2017). Given that neural oscillations are built upon intricate molecular, cellular, and inter-cellular infrastructures and conform to electrophysiological constraints (Hodgkin and Huxley, 1952; FitzHugh, 1961; T. W. Lee, 2016), it is reasonable to assume that injecting artificial alternating currents will force the neural dynamics moving away from its balanced optimal state and is unfavorable for neural synchronization.

Interestingly, the contralateral side of the administered tACS presented with enhanced CSD powers across spectra, again most prominent for beta and gamma, and the trend persisted to the lower frequency range. The bi-hemispheric reverse pattern of power response to tACS is a frequently observed phenomenon in the field of tES research. The most probable neural mechanism to mediate this is through inter-hemispheric rivalry (Kinsbourne, 1974; Hilgetag et al., 2001; Naeser et al., 2005). The induced transient reduction in neural activity in one region will disrupt the dynamic equilibrium, and the ensuing decreased inhibitory output will enhance the neural activity in the homologous area of the contralateral hemisphere. The inverse trend of power changes also contributes to keeping the brain’s energy expenditure relatively stable (T. W. Lee, 2016), in accordance with the concept of "net zero-sum" (Brem et al., 2014).

Notably, our 40-Hz tACS results were highly concordant with our recent work of 5-Hz analog (T. W. Lee and Tramontano, 2024c). We thus conclude that narrow band tACS in fact interferes with underlying neural synchronization across broad spectra (spectrum-unspecific), which in turn augments contralateral oscillatory powers. The neural spectral impact of tACS diminishes as the separation from the default frequency (i.e., 40 Hz in this research) increases. Since entrainment theory is based on electrophysiological research via invasive recording, how to reconcile the two seemingly contradictory observations? In fact, the “entrainment” may not be reflected in the magnitude of power, but in the connectivity strength at the delivered frequency band (spectrum-specific), see below.

### Connectivity changes associated with tACS treatment

After 40-Hz tACS treatment, the lagged phase synchronization at gamma range significantly increased between the frontal and parietal regions. The increase in lagged coherence did not pass statistical challenge. The pattern replicated our recent work of 5-Hz tACS (T. W. Lee and Tramontano, 2024b). Compelling evidence indicates that tACS can modulate the timing of neuronal spiking activity (Krause et al., 2019; Anastassiou et al., 2010; Radman et al., 2007). When tACS is applied to two distinct brain regions, it is anticipated to enhance synchronization of neural firing in these areas, with the timing guided by the applied alternating currents and the corresponding oscillatory frequencies. Mathematically speaking, the formula for phase synchronization closely resembles that of coherence, with the sole distinction being the inclusion of a normalization procedure aimed at mitigating the impact of power. Interestingly, the same as our previous work (T. W. Lee and Tramontano, 2024b), the index based on lagged phase synchronization proved to be more robust than that derived from lagged coherence, indicating that what was “entrained” was not power but phase relationship.

In addition, since the recorded EEGs were spanned by 12 sessions of tACS treatment, not real-time, the significant inter-regional interactions suggest the activation of a plasticity mechanism. Similarly, Vossen and colleagues investigated the after-effects of alpha tACS and claimed that tACS’s modulatory influence was driven by plasticity as opposed to entrainment (Vossen et al., 2015). To conclude, at the large-scale network level, the inter-regional effects of tACS were mediated by phase synchronization (crosstalk) and subsequent involvement of neural plasticity, not by the concurrent entrainment of powers. Our separate tACS studies altogether offer novel insights to clarify the debates surrounding the applicability of entrainment theory. Furthermore, if tACS impairs underlying neural synchronization, the treatment benefits may be mediated by enhanced frontoparietal connectivity strength.

Previous studies investigating the effects of tACS on functional connectivity indices have shown inconsistent patterns, such as the direction of connectivity change and spectral specificity. For example, frontal theta tACS has been reported to enhance phase-locking values between frontal and posterior regions across a broad spectrum (Jones et al., 2022), whereas delta tACS increased phase lag indices in the theta range rather than in delta (Davis et al., 2023). Bilateral gamma tACS was found to decrease inter-hemispheric connectivity, modulated by different phase lags (Preisig et al., 2021). Other studies have shown spectral specificity in the modulation of functional connectivity, indicating that different frequencies can selectively enhance connectivity across various brain networks (Aktürk et al., 2022). Comparative research has raised concerns about the validity of using scalp signals to study cortical connectivity, as cortical sources typically do not project radially to the scalp, which can lead to inaccurate or incomplete representations of brain network interactions (Lehmann et al., 2012; RD Pascual-Marqui et al., 2014). The striking consistency observed in our separate tACS studies underscores the advantages of a brain-based tomography approach compared to scalp-based, topographical methods.

We acknowledge several limitations in this preliminary report, including the potential influence of practice effects (especially neuropsychological tests) and the need for severity criterion to better stratify participants, which could reduce heterogeneity and lead to more precise conclusions. The absence of a sham control group complicates the interpretation of results, making it difficult to distinguish between genuine treatment effects and placebo responses. Although the 10-20 EEG system has a limited number of electrodes, nonetheless, given that both EEG and tACS are low-resolution tools (the latter is evident from simulation computation) (Saturnino et al., 2017; C. Lee et al., 2017), increasing the electrode count may not yield significant advantages.

## Conclusion

This study investigated the neural effects of 40-HZ tACS at 2.0 mA over the left frontoparietal network (F3 and P3) by comparing EEG recordings before and after 12-session treatment. To realize the tomography approach, the EEG registered from the scalp were converted to brain signals using eLORETA. The resulting neural changes were consistent with our previous report of 5-Hz tACS over the right hemisphere. In summary, the neural patterns subsequent to tACS included regional de-synchronization across broad spectra, accompanied by contralateral enhancement of power, and an enhancement of inter-regional phase synchronization at the default frequency. We thus infer that the “entrainment” by tACS may exert its influence on the functional connectivity, not oscillatory power, in post-stimulation conditions.

## Statement of Ethics

### Study approval statement

This research analyzed data collected from 2018 to 2022. This study protocol was reviewed and approved by Pearl IRB (https://www.pearlirb.com/), approval number 2023-0133, and was granted ‘exempt’ status.

### Consent to participate statement

Written informed consent was obtained from all participants involved in this study. For participants under 18 years of age, the written informed consent was provided by their parents, legal guardians, or next of kin.

## Authors Contributions

Both authors contributed intellectually to this work. TW Lee carried out the analysis and wrote the first draft. Both authors revised and approved the final version of the manuscript.

## Data Availability Statement

The data supporting the findings of this study are not publicly available due to restrictions outlined in the informed consent. However, they are available from the corresponding author upon reasonable request.

